# Self-Reported Side Effects Among Reddit Users Taking Nonapproved Retatrutide

**DOI:** 10.64898/2026.05.28.26352819

**Authors:** Neil K. R. Sehgal, Jena Shaw Tronieri, Benjamin Rader, Lyle Ungar, Sharath Chandra Guntuku

## Abstract

Gray-market retatrutide use is increasing, but patient safety experiences remain poorly characterized. This cross-sectional analysis examined Reddit posts and comments from retatrutide-specific and broader peptide or weight-management communities through December 2025. A validated large language model classified self-reported retatrutide use and extracted author-attributed symptoms mapped to MedDRA Preferred Terms. Among 13,589 users reporting current use, 7,823 had at least one mapped symptom after exclusions. Unlike phase 2 trial findings dominated by gastrointestinal events, Reddit reports most often described appetite increase, fatigue, increased energy, nausea, food craving, insomnia, and elevated heart rate. Findings are hypothesis-generating and warrant pharmacovigilance attention.

## Introduction

The gray-market trade in nonapproved peptides has expanded rapidly alongside the rise of glucagon-like peptide 1 receptor agonists (GLP-1 RA), creating a population of patients whose safety experiences remain largely invisible to regulators and clinicians [1]. Retatrutide is an investigational once-weekly agonist of the glucose-dependent insulinotropic polypeptide, GLP-1, and glucagon receptors. The US Food and Drug Administration has warned that nonapproved products containing retatrutide are being sold directly to consumers, including products labeled for research or not for human consumption, yet no systematic characterization of symptoms among real-world, unregulated use exists [2]. Public interest has also expanded rapidly: in U.S. Google Trends data, relative search interest for retatrutide rose from near zero before mid-2023 to over three-quarters of semaglutide search interest by April 2026 [3]. Furthermore, FDA data suggests hundreds of thousands of doses entered the US between January and April 2026 [4]. In a phase 2 obesity trial, gastrointestinal (nausea, diarrhea, vomiting, and constipation) adverse events were most common, with phase 3 trials ongoing [5].

Social media can complement traditional pharmacovigilance approaches by offering insight into patients’ experiences that may not be fully represented through traditional spontaneous reporting systems [6]. This may be particularly relevant for retatrutide, for which the total number of treated participants in published clinical trials is small, while individuals obtaining the drug through unregulated channels outside clinical trials may not be captured by conventional pharmacovigilance systems. The large volume and rapid availability of user-generated content may facilitate more timely monitoring of emerging or previously unidentified safety signals when appropriately analyzed [7]. Reddit is particularly suited for this purpose due to its large, publicly available, topic-specific health communities and its long-form, context-rich user discussions.

Prior work has established Reddit as a valuable source for studying health-related disclosures, including sensitive or stigmatized medical experiences [8, 9]. Importantly, such observations are hypothesis-generating and require independent validation rather than being interpreted as estimates of incidence or evidence of causality.

We sought to characterize self-reported symptoms among Reddit users describing personal retatrutide use, including discussions likely to reflect nontrial use of unregulated, nonapproved products, to identify potential safety signals warranting formal pharmacovigilance attention.

## Methods

We conducted a cross-sectional analysis of Reddit posts and comments from 6 retatrutide-specific and 21 broader peptide or weight-management communities, collected May 1, 2021, through December 31, 2025. After applying prespecified exclusion criteria (e.g., removing common bot accounts, posts with fewer than 10 words, etc.), we retained 148,640 retatrutide-related posts or comments from 38,936 unique users.

A large language model (gpt-5.4-nano) classified author self-disclosed retatrutide use and extracted disclosed dose and demographic details. A second model extracted author-attributed symptoms and mapped them to Medical Dictionary for Regulatory Activities (MedDRA) Preferred Terms (PTs). After excluding prespecified weight-loss and appetite-suppression PTs, PTs were aggregated at the user level, mapped to primary System Organ Class and High-Level Terms (HLT), and reported descriptively. Manual validation of 100 random posts showed high precision and recall for use classification and symptom extraction. For context, HLT frequencies from prior Reddit analyses of semaglutide/tirzepatide were reproduced; no statistical comparisons were performed [10]. All analyses were conducted in Python 3.11. Full methodological details are provided in the Supplement. As the data were public and non-identifiable, institutional review board review was not required. Strengthening the Reporting of Observational Studies in Epidemiology (STROBE) reporting guidelines were followed.

## Results

Of 148,640 retatrutide-related posts/comments, the classifier identified 45,592 posts/comments from 13,589 unique users reporting current retatrutide use, dated March 10, 2023, through December 31, 2025. Among these users, 9699 (71.4%) had at least 1 extracted MedDRA concept before mapping and exclusions. After excluding prespecified weight-related and appetite-suppression PTs, 7823 users (57.6%) had at least 1 mapped PT and formed the analytic denominator for symptom frequencies.

The most common PTs were increased appetite, fatigue, increased energy, nausea, food craving, insomnia, and heart rate increased (Table 1). In a descriptive comparison with HLT frequencies reproduced from a prior semaglutide/tirzepatide Reddit study, retatrutide discussions were more concentrated in appetite, general-system, muscle-related, sleep, and heart-rate/pulse categories, whereas semaglutide/tirzepatide discussions were more concentrated in nausea/vomiting and gastrointestinal motility categories (Figure 1).

**Figure 1.**
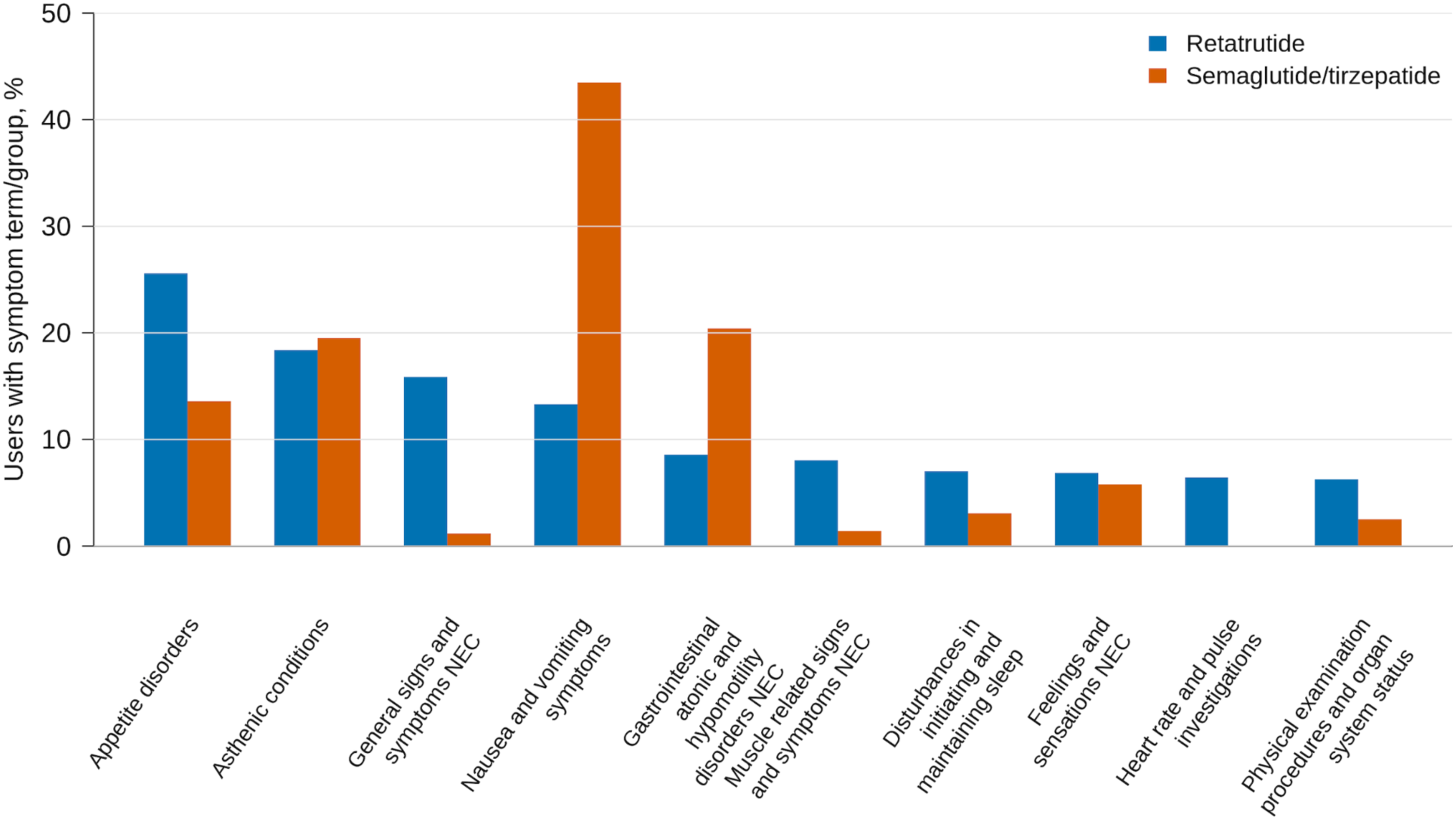
MedDRA HLT Symptom Groups Reported by Retatrutide and Semaglutide/Tirzepatide Reddit Users. Bars show user-level percentages for the 10 most frequent retatrutide MedDRA High-Level Terms after combining appetite-related PTs. Retatrutide values are from the current study. Semaglutide/tirzepatide values are reproduced from prior work [10] and are shown for descriptive context only. Samples were not matched on demographic or clinical characteristics, and no statistical comparisons were performed.

**Table 1.** MedDRA Preferred Terms Reported by Reddit Users Self-Reporting Retatrutide Use. PTs reported by less than 0.5% of users and System Organ Classes reported by less than 1.0% of users were omitted for brevity. Prespecified weight-loss-related and appetite-suppression PTs were excluded before calculating the denominator. SOC rows count users with at least 1 PT in that SOC, so counts do not sum across rows.

| MedDRA primary System Organ Class and Preferred Term | No. (%) of users (n=7823) |
| --- | --- |
| **General disorders and administration site conditions** | **3396 (43.4)** |
| Fatigue | 1188 (15.2) |
| Energy increased | 970 (12.4) |
| Hunger | 366 (4.7) |
| Malaise | 318 (4.1) |
| Inflammation | 208 (2.7) |
| Pain | 148 (1.9) |
| Drug ineffective | 144 (1.8) |
| Feeling abnormal | 130 (1.7) |
| Exercise tolerance increased | 130 (1.7) |
| Feeling cold | 120 (1.5) |
| Exercise tolerance decreased | 94 (1.2) |
| Injection site pain | 85 (1.1) |
| Chills | 74 (0.9) |
| Asthenia | 58 (0.7) |
| Thirst decreased | 56 (0.7) |
| Therapeutic product effect decreased | 48 (0.6) |
| Oedema | 45 (0.6) |
| Feeling hot | 44 (0.6) |
| Hangover | 44 (0.6) |
| Insulin sparing effect | 40 (0.5) |
| Temperature intolerance | 40 (0.5) |
| **Metabolism and nutrition disorders** | **2578 (33.0)** |
| Increased appetite | 1336 (17.1) |
| Food craving | 571 (7.3) |
| Abnormal weight gain | 186 (2.4) |
| Hypoglycaemia | 163 (2.1) |
| Food aversion | 143 (1.8) |
| Dehydration | 143 (1.8) |
| Hyperphagia | 120 (1.5) |
| Lack of satiety | 106 (1.4) |
| Hypermetabolism | 88 (1.1) |
| Fluid retention | 71 (0.9) |
| Insulin resistance | 68 (0.9) |
| Body fat disorder | 64 (0.8) |
| Weight fluctuation | 61 (0.8) |
| Alcohol intolerance | 58 (0.7) |
| Polydipsia | 42 (0.5) |
| **Gastrointestinal disorders** | **2542 (32.5)** |
| Nausea | 907 (11.6) |
| Constipation | 354 (4.5) |
| Diarrhoea | 345 (4.4) |
| Abdominal distension | 331 (4.2) |
| Abdominal pain | 224 (2.9) |
| Eructation | 218 (2.8) |
| Gastrooesophageal reflux disease | 215 (2.8) |
| Vomiting | 206 (2.6) |
| Dyspepsia | 174 (2.2) |
| Abdominal discomfort | 166 (2.1) |
| Gastrointestinal disorder | 154 (2.0) |
| Impaired gastric emptying | 124 (1.6) |
| Flatulence | 102 (1.3) |
| Dry mouth | 92 (1.2) |
| Abnormal faeces | 78 (1.0) |
| Faeces soft | 53 (0.7) |
| Frequent bowel movements | 49 (0.6) |
| **Psychiatric disorders** | **2367 (30.3)** |
| Insomnia | 475 (6.1) |
| Anxiety | 301 (3.8) |
| Anhedonia | 206 (2.6) |
| Euphoric mood | 188 (2.4) |
| Binge eating | 140 (1.8) |
| Libido decreased | 134 (1.7) |
| Apathy | 111 (1.4) |
| Mood altered | 105 (1.3) |
| Sleep disorder | 101 (1.3) |
| Abnormal dreams | 78 (1.0) |
| Irritability | 75 (1.0) |
| Libido increased | 70 (0.9) |
| Drug dependence | 69 (0.9) |
| Depression | 68 (0.9) |
| Alcoholism | 66 (0.8) |
| Depressed mood | 65 (0.8) |
| Attention deficit hyperactivity disorder | 64 (0.8) |
| Decreased interest | 64 (0.8) |
| Panic attack | 64 (0.8) |
| Restlessness | 59 (0.8) |
| Poor quality sleep | 52 (0.7) |
| Hyposomnia | 47 (0.6) |
| Substance dependence | 43 (0.5) |
| Nicotine dependence | 41 (0.5) |
| Obsessive thoughts | 41 (0.5) |
| Emotional distress | 40 (0.5) |
| Anorexia and bulimia syndrome | 40 (0.5) |
| **Investigations** | **1534 (19.6)** |
| Heart rate increased | 374 (4.8) |
| Weight increased | 305 (3.9) |
| Blood glucose decreased | 165 (2.1) |
| Blood pressure decreased | 126 (1.6) |
| Heart rate | 83 (1.1) |
| Blood cholesterol decreased | 61 (0.8) |
| Blood glucose increased | 56 (0.7) |
| Heart rate variability decreased | 49 (0.6) |
| Blood glucose normal | 45 (0.6) |
| **Nervous system disorders** | **1513 (19.3)** |
| Headache | 237 (3.0) |
| Dizziness | 177 (2.3) |
| Allodynia | 162 (2.1) |
| Hyperaesthesia | 161 (2.1) |
| Dysgeusia | 127 (1.6) |
| Somnolence | 122 (1.6) |
| Paraesthesia | 106 (1.4) |
| Lethargy | 88 (1.1) |
| Brain fog | 73 (0.9) |
| Hypersomnia | 67 (0.8) |
| Cognitive disorder | 65 (0.8) |
| Burning sensation | 56 (0.7) |
| Dysaesthesia | 54 (0.7) |
| Presyncope | 45 (0.6) |
| **Musculoskeletal and connective tissue disorders** | **1125 (14.4)** |
| Muscle hypertrophy | 317 (4.1) |
| Myalgia | 167 (2.1) |
| Muscle atrophy | 153 (2.0) |
| Muscular weakness | 143 (1.8) |
| Arthralgia | 129 (1.6) |
| Musculoskeletal pain | 111 (1.4) |
| Muscle fatigue | 100 (1.3) |
| Muscle mass | 59 (0.8) |
| Back pain | 45 (0.6) |
| Pain in extremity | 40 (0.5) |
| **Skin and subcutaneous tissue disorders** | **776 (9.9)** |
| Sensitive skin | 110 (1.4) |
| Pruritus | 86 (1.1) |
| Hyperhidrosis | 73 (0.9) |
| Skin laxity | 61 (0.8) |
| Skin burning sensation | 55 (0.7) |
| Alopecia | 50 (0.6) |
| Skin sensitisation | 48 (0.6) |
| Night sweats | 41 (0.5) |
| **Cardiac disorders** | **465 (5.9)** |
| Tachycardia | 294 (3.8) |
| Palpitations | 146 (1.9) |
| **Vascular disorders** | **199 (2.5)** |
| Hypotension | 64 (0.8) |
| Hot flush | 57 (0.7) |
| **Social circumstances** | **174 (2.2)** |
| Alcohol use | 60 (0.8) |
| **Renal and urinary disorders** | **139 (1.8)** |
| Nocturia | 53 (0.7) |
| **Immune system disorders** | **113 (1.4)** |
| Hypersensitivity | 96 (1.2) |
| **Eye disorders** | **109 (1.4)** |
| Vision blurred | 44 (0.6) |

## Discussion

In this analysis of Reddit users self-reporting retatrutide use, gastrointestinal symptoms were common but were not the dominant symptom category, a pattern that diverges from its phase 2 trial profile and that of other FDA-approved GLP-1 RAs [5]. Instead, appetite increases, fatigue, increased energy, insomnia, heart-rate terms, and muscle-related symptoms were among the most frequently reported adverse experiences. The unique adverse events identified may be attributable to retatrutide’s glucagon receptor activity as the only tri-agonist in its class. For example, single GLP-1R and dual GIP and GCGR agonists have differed in effects on energy expenditure in rodents, suggesting different metabolic effects [11]. However, frequent reports of increased appetite and hunger were unexpected in light of the 24-week mean body weight losses of up to 17.5% with retatrutide (12 mg/wk) in initial trials [5].

Although self-reported experiences with retatrutide appeared to differ from those of Reddit users taking semaglutide and tirzepatide, such patterns may be attributable to unmeasured cohort differences rather than to the medications’ pharmacology [10]. Additionally, reported events may be attributable to product quality and dose variability, stacking with other peptides (concurrent use of multiple nonapproved peptides, a common practice in online communities), user profile (e.g., popularity among weightlifters), lyophilized peptide reconstitution process, or misattribution, rather than causal drug effects.

These findings should be viewed as hypothesis-generating. Reddit users may not represent all retatrutide users, and users with adverse events may be more likely to post. Further, while some users may have been clinical-trial participants, users obtaining retatrutide through gray-market channels are likely to differ from those enrolled in or eligible for retatrutide obesity and diabetes clinical trials. These differences limit comparisons with clinical-trial populations but may also give rise to safety concerns specific to unregulated use that would not necessarily be captured in clinical trials. In addition, product identity, purity, dose, route, treatment duration, concurrent drugs, and medical history cannot be determined. The analysis cannot estimate incidence, causality, dose-response, or comparative safety vs approved GLP-1 receptor agonists. Natural language processing and MedDRA mapping may also miss or misclassify symptoms, however manual validation suggests performance comparable or superior to past work [10, 12–13].

As gray-market retatrutide use expands, clinicians are likely to encounter patients obtaining this compound outside regulated prescribing pathways. Nonjudgmental screening for nonapproved peptide use, counseling about unknown product quality and the absence of safety data in this population, and reporting of adverse events to MedWatch may improve surveillance and patient safety. Prospective studies are needed to determine whether these signals represent true pharmacological effects and at what doses and durations.

## Data Sharing Statement

Code to reproduce the analyses can be found at https://github.com/sehgal-neil/retatrutide_side_effect_analysis

All data used in this study are publicly available from Reddit.com, or are accessible via Pushshift and Arctic Shift. Due to Reddit’s Terms of Use, we are unable to share the raw data. All Reddit data in this study were used in accordance with Reddit’s Terms of Use.

## Funding/Support

No funding was received for this work.

## Conflict of Interest Disclosures

JST reports receiving an investigator-initiated grant, on behalf of the University of Pennsylvania, from Novo Nordisk and consulting fees from Currax Pharmaceuticals, LLC. No other disclosures were reported.

## Data Availability

All data used in this study are publicly available from Reddit.com or are accessible via Pushshift and Arctic Shift. Due to Reddits Terms of Use we are unnable to share the raw data. All Reddit data in this study were used in accordance of Reddits Terms of Use. As data were public and non-identifiable IRB review was not required.

## SUPPLEMENTARY MATERIALS

### Data Source

Reddit posts and comments were collected from Reddit data derived from Pushshift and Arctic Shift. The collection period was May 1, 2021, through December 31, 2025. This overall query window was selected to include early retatrutide discussion (the first phase 2 trial began May 2021) [5]. While the retatrutide-related dataset spanned May 1, 2021 through December 31, 2025, the first post/comment retatrutide self-use was dated March 10, 2023. The archive included public submissions and comments only. No private, password-protected, or deleted content was accessed beyond what existed in the public archive. As data were public and non-identifiable, institutional review board review was not required.

### Subreddit Sampling

Two tiers of subreddits were searched.

Subreddits included both retatrutide-specific communities and broader peptide, exercise and weight-management, and glucagon-like peptide-1 related communities. Communities were selected to maximize sensitivity for retatrutide discussion. Retatrutide-specific communities were included to capture concentrated retatrutide discussions, and broader peptide, glucagon-like peptide 1, exercise, and weight-management communities were included because retatrutide was discussed in contexts involving peptide sourcing, stacking, body composition, and weight management. This was not a probability sample of Reddit communities or retatrutide users and should not be interpreted as representative of all people using retatrutide.

These included: “RetatrutideGBP”, “Retatrutide”, “RetatrutideWomen”, “RetatrutideTrial”, “retatrutide4obesity”, “Retatrutidevendors”, “Biohackers”, “Zepbound”, “GymMotivation”, “WeightLossAdvice”, “BariatricSurgery”, “workout”, “fit”, “Mounjaro”, “Peptidesource”, “BodyHackGuide”, “Peptides”, “PeptideGuidesPH”, “compoundedtirzepatide”, “peptidess”, “PeptidePathways”, “USPeptides”, “PeptideGuide”, “PeptideDiscussion”, “PeptideForum”, “PeptideProgress”, and “tirzepatidecompound”

### Keyword Strategy

The following retatrutide names, abbreviations, and common misspellings were used to filter for posts mention retatrutide: “retatrutide”, “reta”, “retatrutid”, “retaturtide”, “retratrutide”, “retatritide”, “retatruide”, “retartutide”, “retatrutyde”, “reterutide”, “retatrudite”, “retatruitde”, “retratutide”, “retatrutid”, “retatatrutide”, “retatrutiede”, and “retatratide”.

### Content Filtering

Comments and submissions were excluded if the author identifier matched a common bot account (e.g., Automoderator, Shakespearebot), if the text was missing, if the text was marked as removed or deleted, if the post/comment had fewer than 10 words, if it lacked spaces in the first 2048 characters, or if fewer than 50% of characters were alphabetic.

### Self-Use Classification

Self-use classification used the OpenAI model `gpt-5.4-nano-2026-03-17`. The classifier was instructed to identify only self-disclosures of retatrutide use by the author. Manual review of 100 random entries found 94.4% precision and 97.1% recall for self-use classification.

### Full Self-Use Classification Prompt

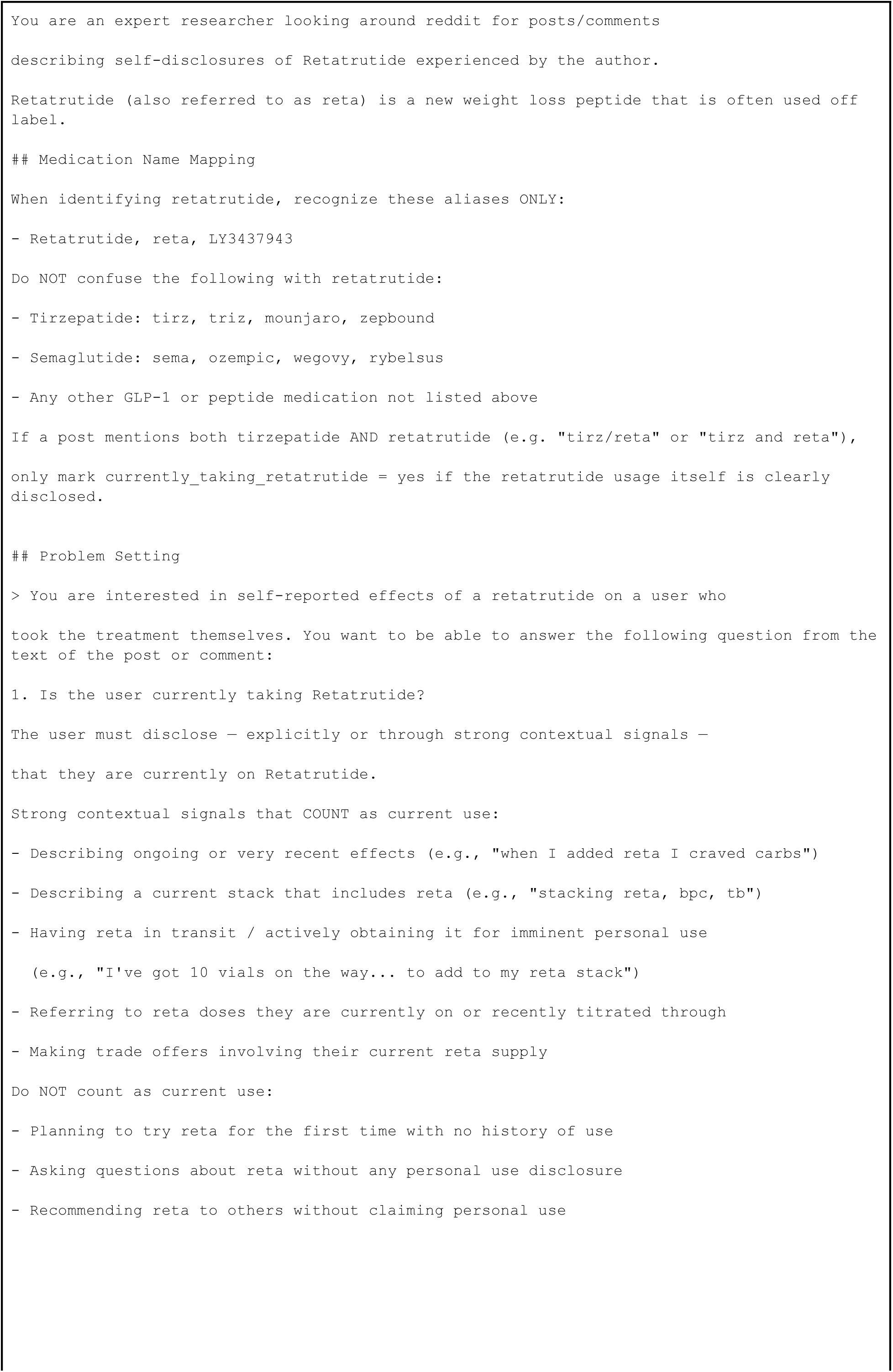
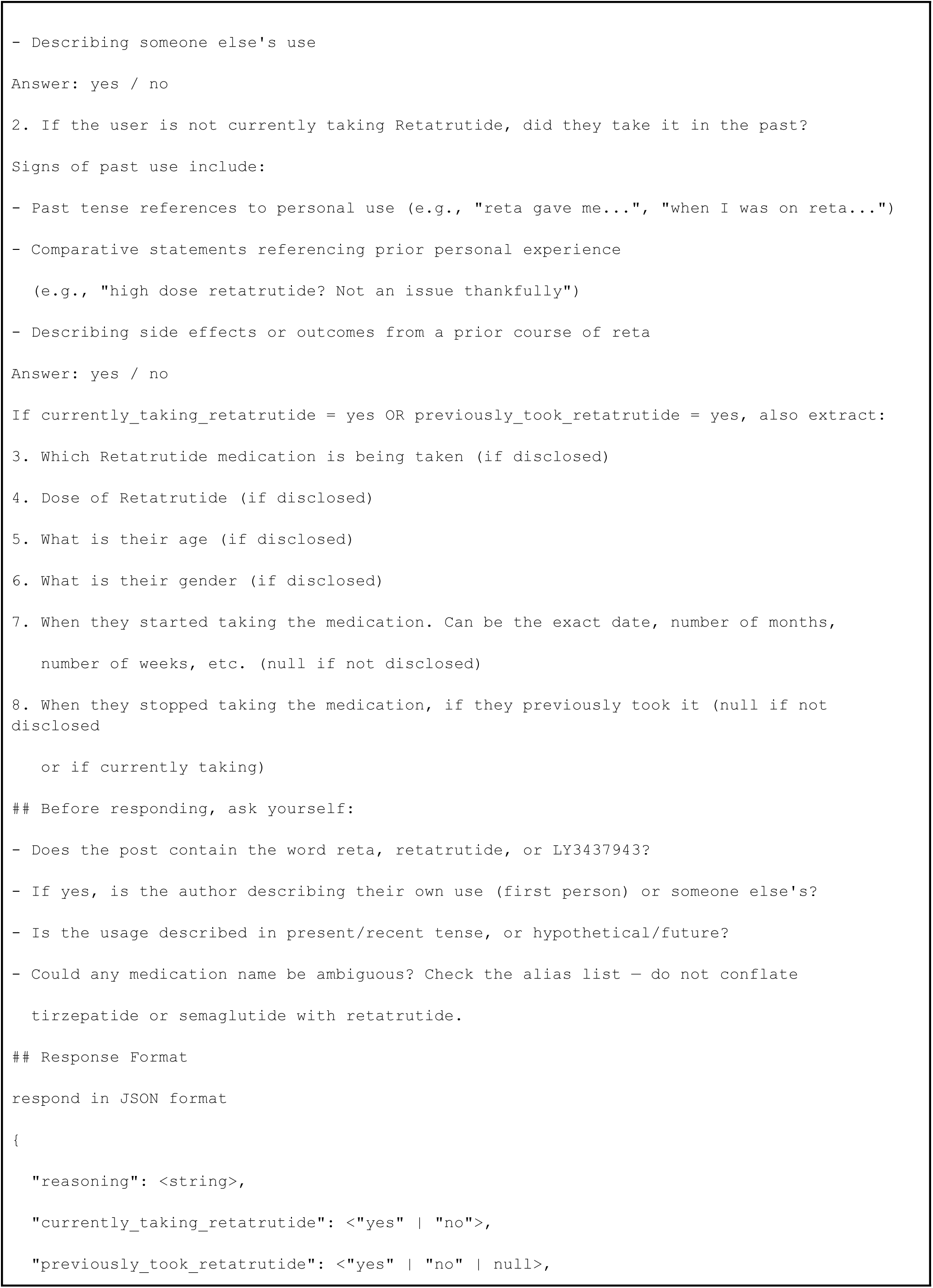
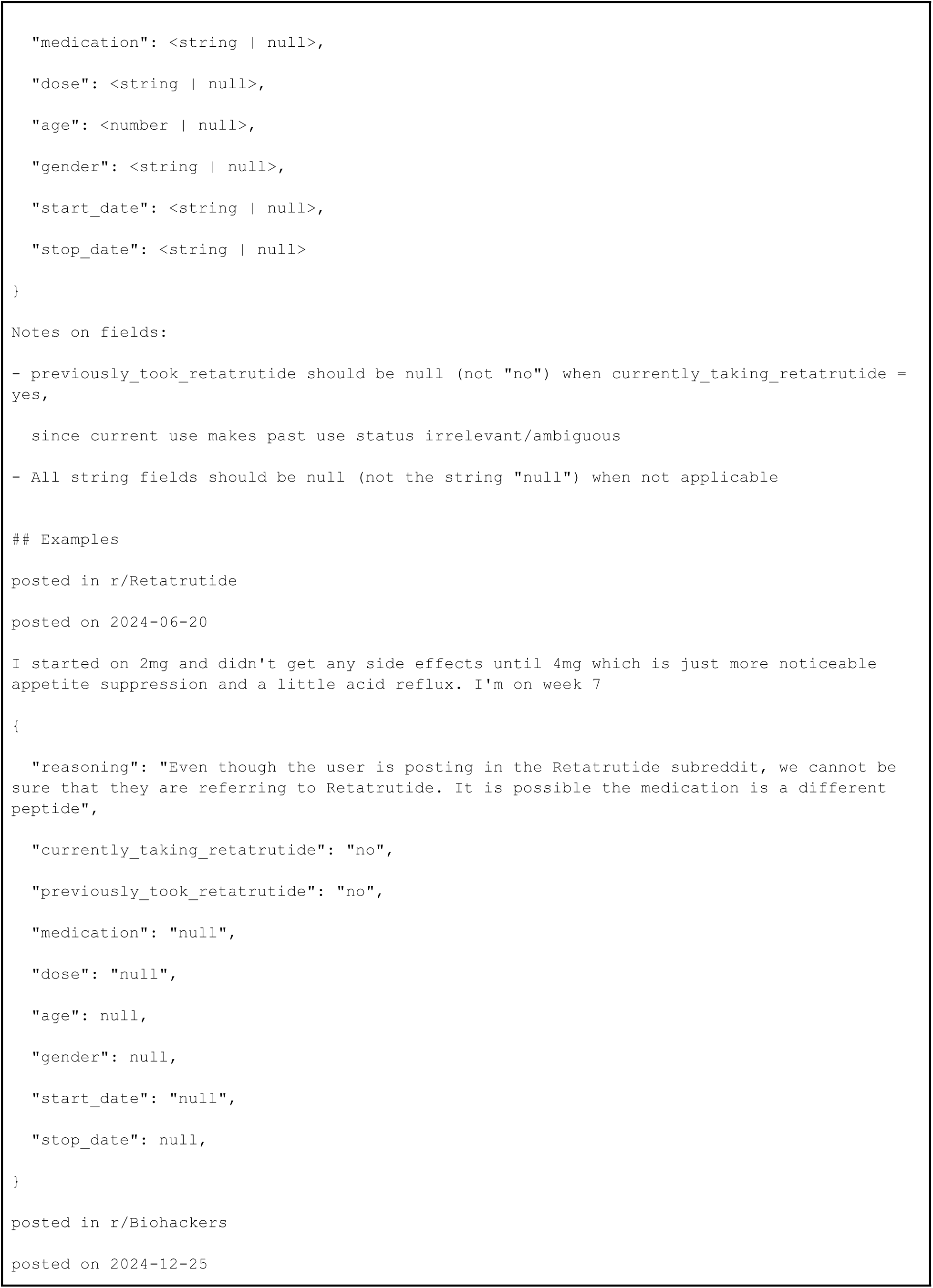
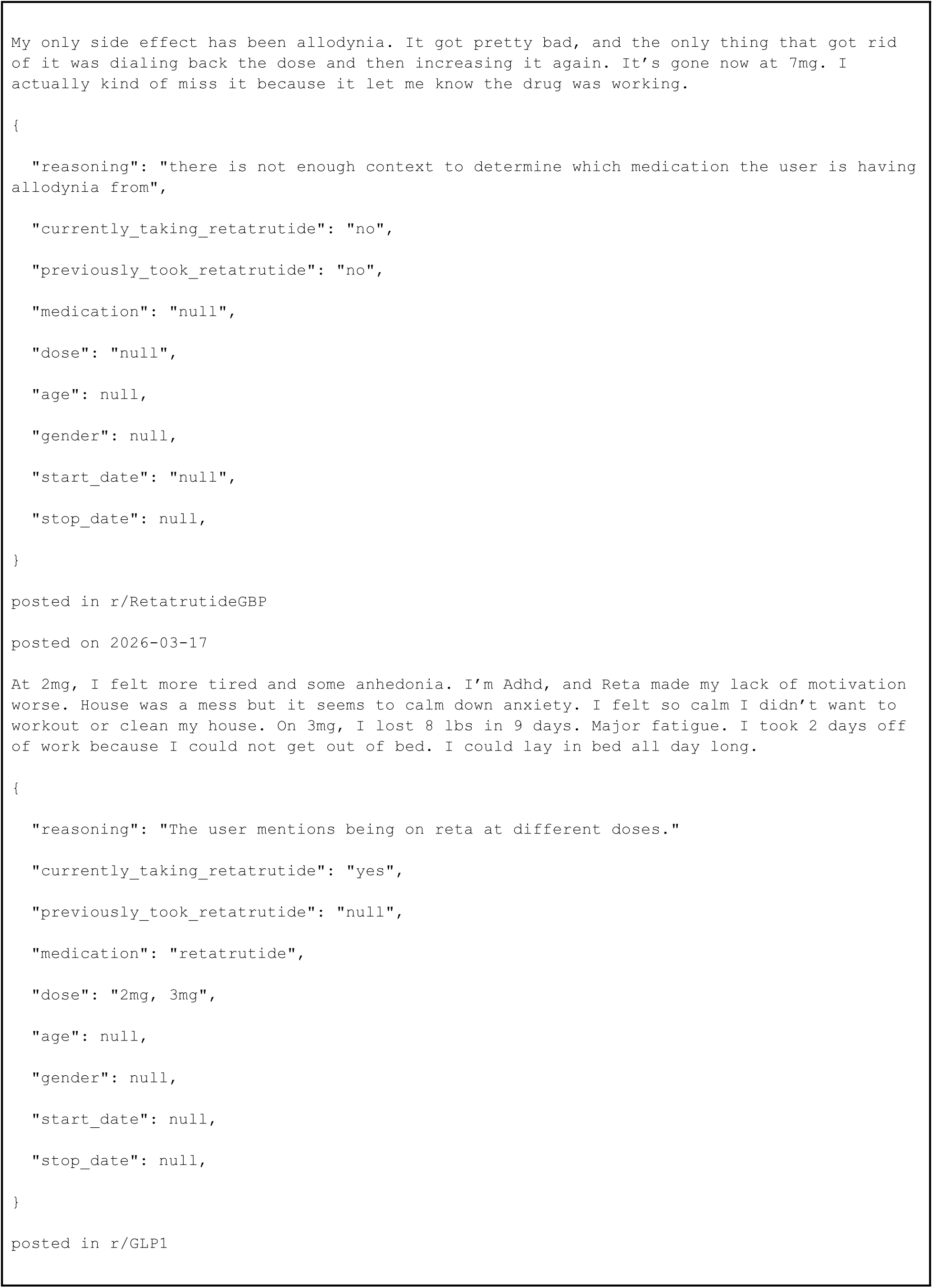
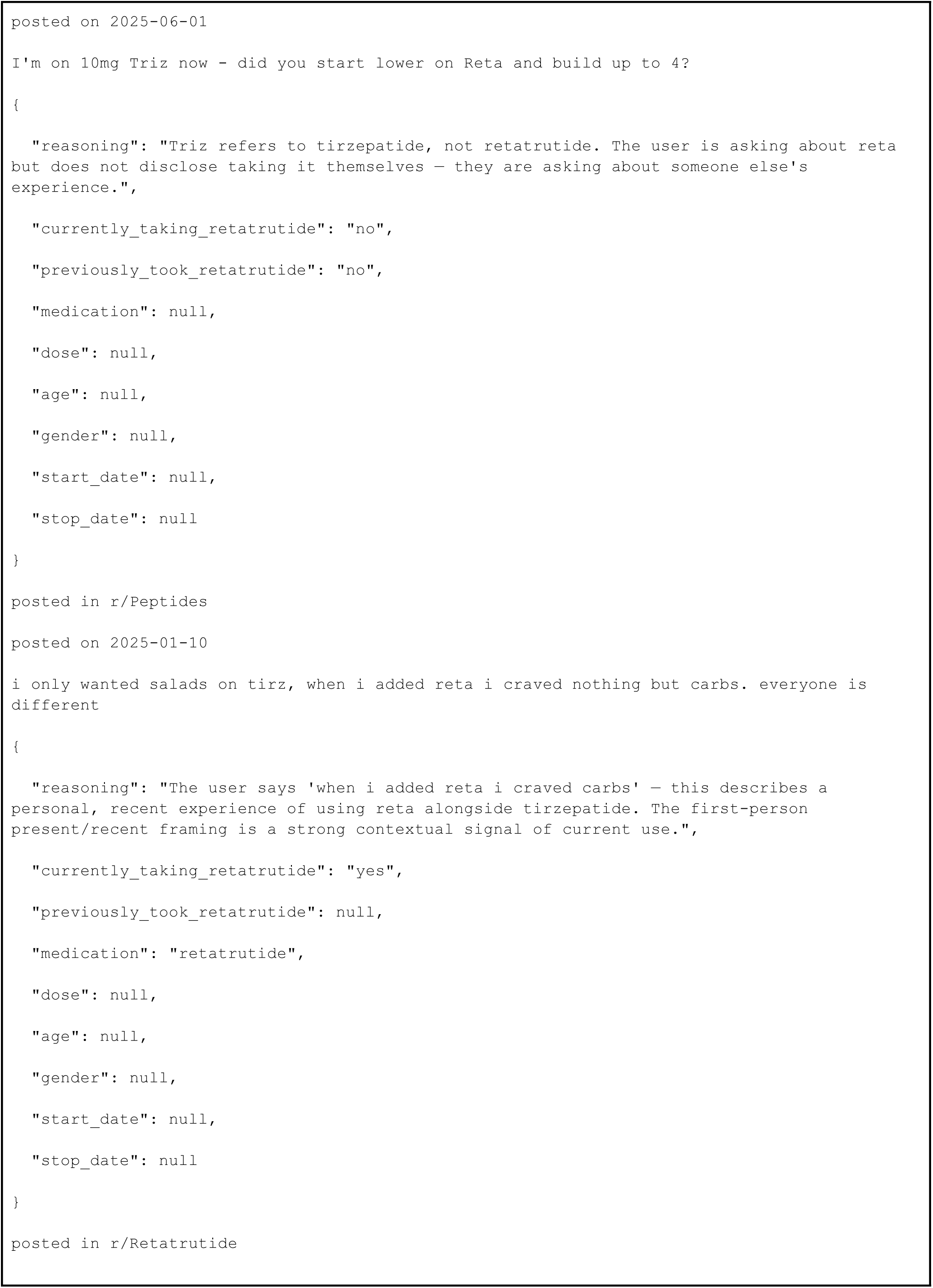
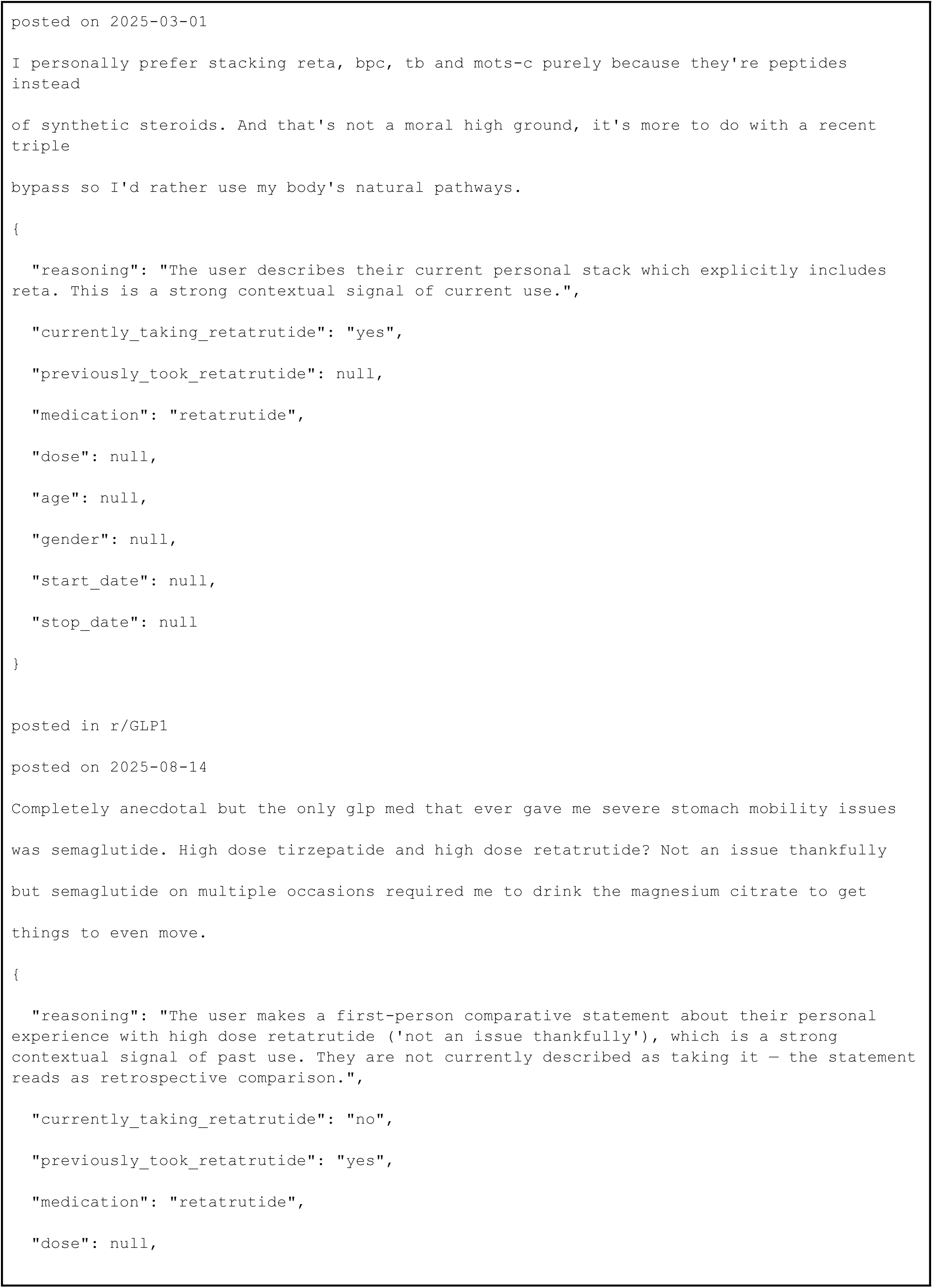
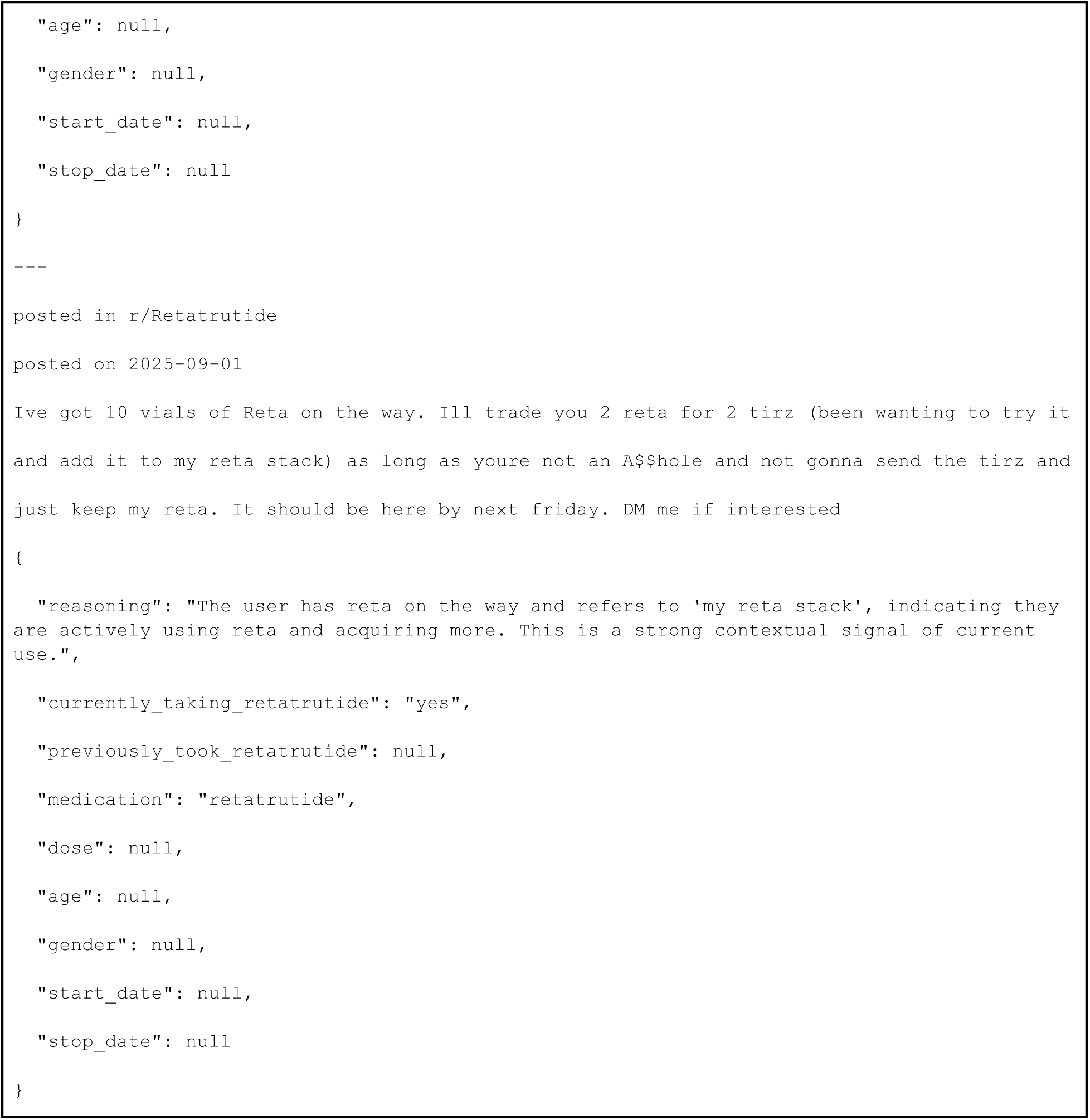

### Side-Effect Extraction

Side-effect extraction used the OpenAI model `gpt-5.4-nano-2026-03-17` with retrieval augmented generation to map side effects to MedDRA Preferred Terms. After excluding prespecified weight-related and appetite-suppression terms, manual review found 91.0% positive predictive value and 98.6% recall for extracted side effects on a random sample of 100 self-use classified posts.

### Full Side-Effect Extraction Prompt

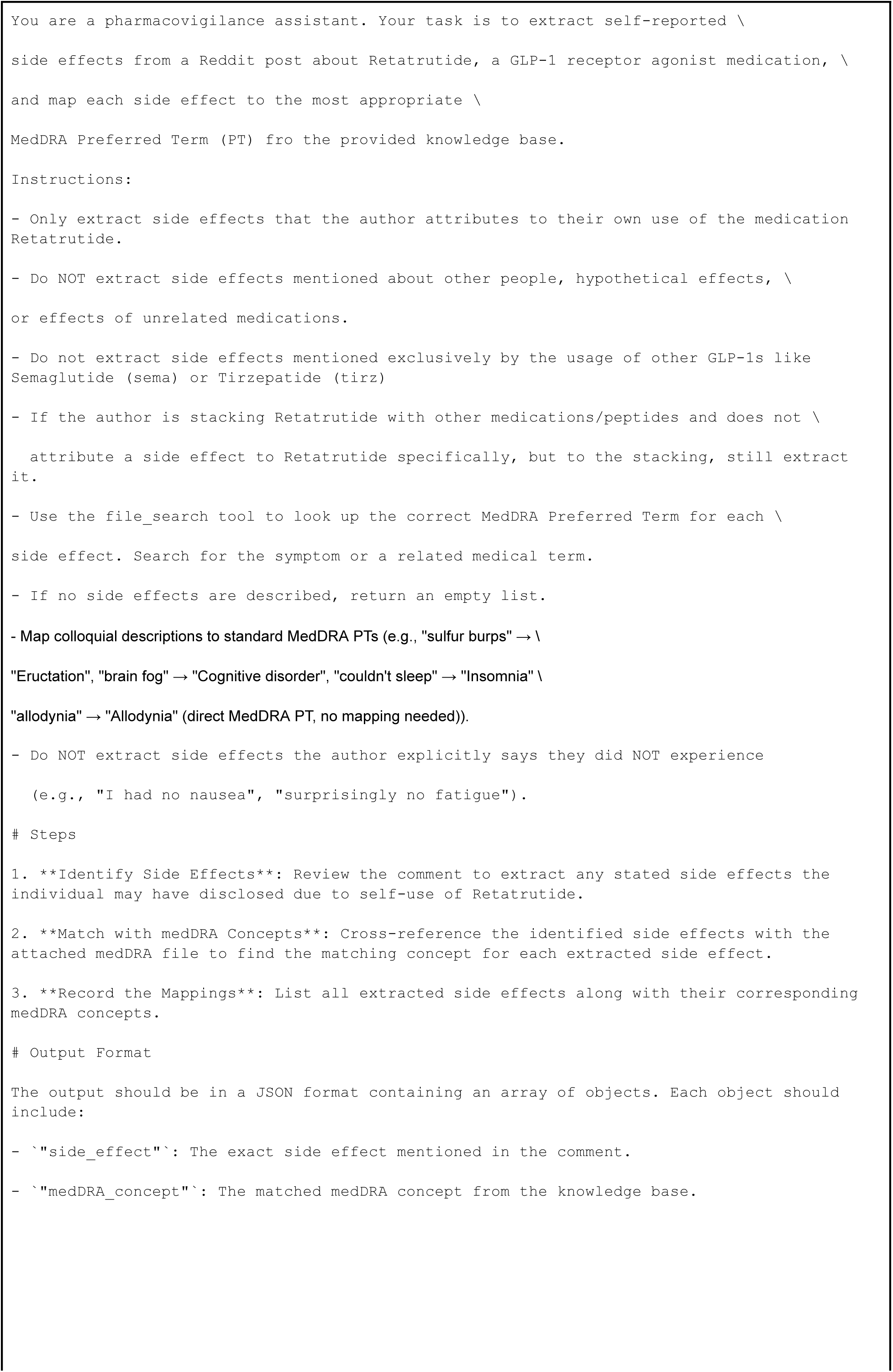
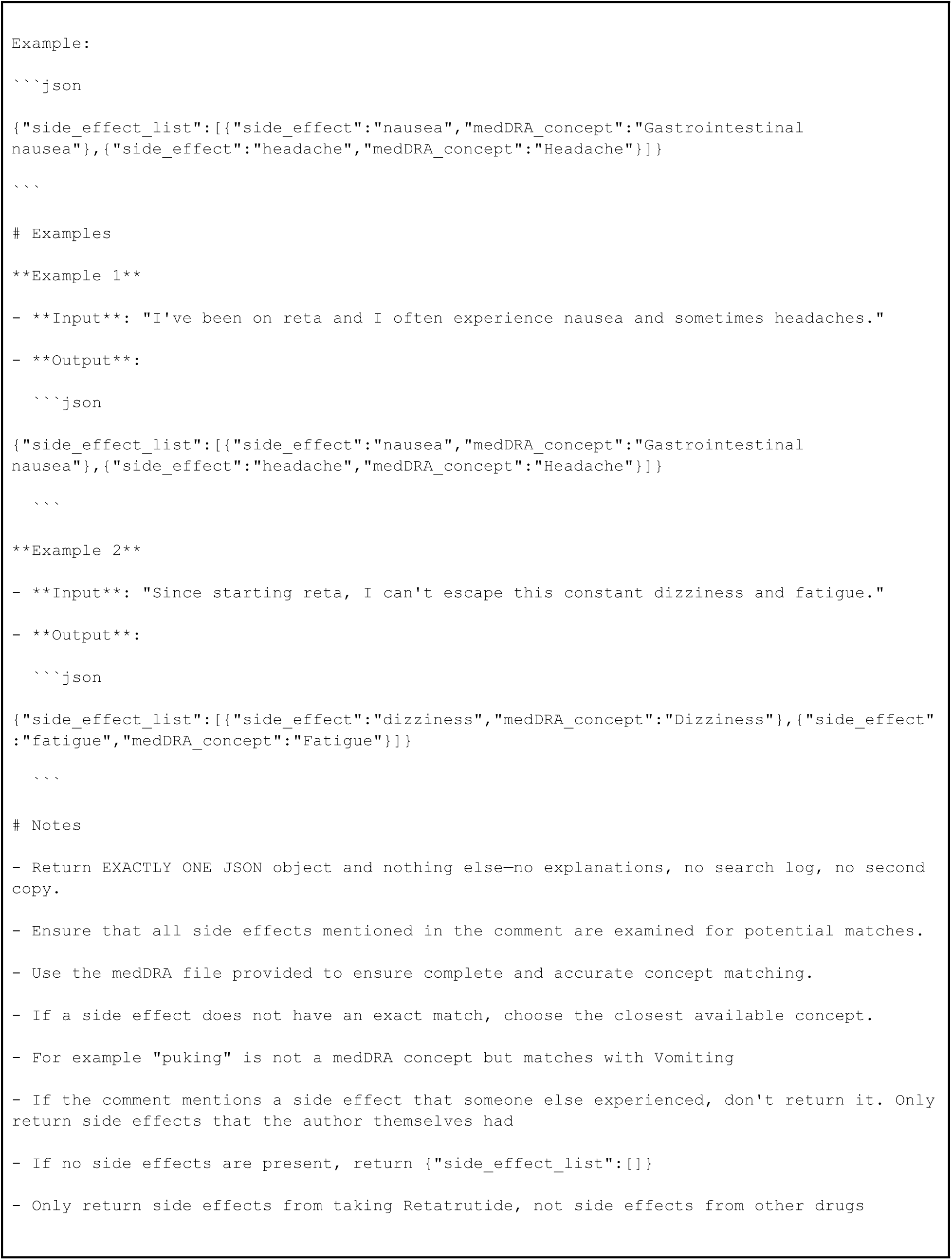

### MedDRA Mapping and Aggregation

Extracted MedDRA concept strings were mapped to official MedDRA PT codes using three steps: exact PT name match, Lower Level Term to parent PT mapping, and manual mapping for any remaining unmatched strings.

Posts were aggregated to the user level by taking the union of all mapped PT codes across that user’s posts/comments. PTs were mapped to High-Level Terms, High-Level Group Terms, and primary System Organ Classes using the MedDRA hierarchy. Users with no remaining mapped PTs after exclusions were omitted from symptom-frequency denominators.

The following weight-related and appetite-suppression PTs were excluded before user-level frequency calculation:

| Excluded PT |
| --- |
| Weight decreased |
| Abnormal loss of weight |
| Decreased appetite |
| Hypophagia |
| Appetite disorder |
| Early satiety |
| Fat tissue decreased |
| Weight loss poor |
| Waist circumference decreased |
| Weight control |

The exclusion was intended to remove weight-related outcomes, weight-control measures, and appetite-suppression terms from the safety-signal tables. Appetite-increase terms, food craving, weight gain terms, and other symptoms were retained.

## Notes

### Summary of Updates

Revised for clarity. No new data or analyses

